# Serological prevalence of SARS-CoV-2 antibody among children and young age (between age 2-17 years) group in India: An interim result from a large multi-centric population-based seroepidemiological study

**DOI:** 10.1101/2021.06.15.21258880

**Authors:** Puneet Misra, Shashi Kant, Randeep Guleria, Sanjay K Rai, WHO Unity Seroprevalence study team of AIIMS

## Abstract

**Background:** Concern has been raised in India regarding the probable third wave of COVID-19 where children and young age group is thought to get affected the most. There is a lack of serological prevalence data in this age group. We have some interim data from our research for WHO unity protocol, which might help policymakers and the research community to answer such questions based on evidence. Hence, we conducted a study to compare the COVID -19 sero-positivity rate between children and adults

**Methods/Materials:** This is part of an ongoing large multi-centric population-based sero-epidemiological study. The study is being conducted in five selected states with a proposed total sample size of 10,000. We have data of 4,500 participants at the time of midterm analysis from four states of India. Total serum antibody against SARS-CoV-2 virus was assessed qualitatively by using a standard ELISA kit. Here we are reporting the interim data of serological prevalence among children aged between 2 to 17 years along with a comparison with ≥18-year old participants.

**Results:** The data collection period was from 15^th^ March 2021 to 10^th^ June 2021. Total available data was of 4,509 participants out of which <18 years were 700 and ≥18 years was 3,809. The site-wise number of available data among the 2-17 year age group were 92, 189, 165, 146 and 108 for the site of Delhi urban resettlement colony, Delhi rural (Villages in Faridabad district under Delhi NCR), Bhubaneswar rural, Gorakhpur rural and Agartala rural area respectively. The seroprevalence was 55.7% in the <18 years age group and 63.5% in the ≥ 18 year age group. There was no statistically significant difference in prevalence between adult and children.

**Conclusion:** SARS-CoV-2 sero-positivity rate among children was high and were comparable to the adult population. Hence, it is unlikely that any future third wave by prevailing COVID-19 variant would disproportionately affect children two years or older.

## Introduction

After the introduction of an infectious agent, the progression of the disease is to a large extent determined by host factors. The age of the host, along with many other factors e.g. pre-existing immunity, co-morbidities etc. plays an important role in determining the further course of the event.^[1]^

Children, particularly aged 5-18 years attend schools. It is commonly believed that classrooms could become outbreak cluster.^[2]^ It is further assumed that those children could then bring the infection home and pass it on to their elderly grandparents who are at a higher risk of dying due to COVID-19. Because of their reasoning, globally there has been occurrence of closure of schools and thereby disadvantaging children in receiving education.^[3,4]^

This line of reasoning presumes that children have a much lower rate of infection as compared to the community at large.^[5,6]^ However, strong evidence is lacking to support this assumption.

We, therefore, undertook a community-based sero-survey for COVID-19 among a population older than two years. The objective of the study was to compare the COVID-19 sero-positivity rate between children and adults.

## Methodology

### Design and study setting

This is part of an ongoing multi-centric population-based, age-stratified prospective COVID-19 sero-prevalence study under WHO (World Health Organisation) Unity studies.^[7]^ It is being conducted in five selected sites in India. The sites are AIIMS (All India Institute of Medical Sciences), New Delhi; AIIMS, Bhubaneswar, Odisha; AIIMS, Gorakhpur, Uttar Pradesh; JIPMER (Jawaharlal Institute of Postgraduate Medical Education & Research), Puducherry and Agartala Medical College, Tripura. In each site both urban and rural area population have been planned to be included. In Delhi, the urban area was a resettlement colony in the South Delhi district where the majority of the population were from lower socio-economic strata. The rural site for Delhi was in the villages of Ballabgarh block in Faridabad district of Haryana which comes under Delhi NCR (National Capital Region). AIIMS Bhubaneswar area was situated in Bhubaneswar, the state capital of Odisha. The AIIMS Gorakhpur area was near the city Gorakhpur, Uttar Pradesh which is a transit point of international surface transport to Nepal. The Agartala site was in the north-eastern state of Tripura. The JIPMER, Puducherry site was a Union Territory situated in South India. In each study site, all family members of ≥2 years of age from at least 10 consecutive families were included from each of the 25 randomly selected clusters assuming four participants per family, if we could not get 40 participants in 10 houses we kept on going until sample of 40 in that cluster was achieved. Thus, we would achieve enrolment of 1,000 participants in each of the urban and rural sites separately. So in each site, the proposed sample size is 2,000 aggregating a total of 10,000 in all five sites. The study was approved by the Institutional ethics committee of AIIMS, New Delhi and all other participating institutions.

Currently, we are reporting the available data of 2-17 years age group from Delhi urban and rural sites of, National Capital region of Delhi (NCR), Bhubaneswar, Gorakhpur and Tripura. The data was collected during 15^th^–31^st^ March 2021 in Delhi urban site, 12^th^ April-22^nd^ May 2021 in Delhi rural (NCR), 22^nd^ March-7^th^ May 2021 in Bhubaneswar rural, 26^th^ March-1^st^ June 2021 in Tripura. Informed consent was obtained from all the participants and the samples were collected maintaining all recommended SOPs.

The primary outcome variable was the participants’ serum antibody reactivity to SARS-CoV-2 virus. Whereas exposure variables were basic demographic variables, clinical variables related to symptoms etc. Five ml venous blood sample was collected in the yellow top plain vial with gel separator from each participant. The total antibodies in serum to SARS-CoV2 were assessed using an Enzyme-linked Immunoassay (ELISA) (Kit: WANTAI SARS-CoV-2 Ab ELISA kit, Wantai SARS-CoV-2 Diagnostics) as per the manufacturer’s protocol. WANTAI SARS-CoV-2 Ab ELISA is an enzyme-linked immunosorbent assay (ELISA) for the qualitative detection of total antibodies against S-RBD SARS-CoV-2 virus in human serum or plasma specimens. It has a sensitivity of 94.4% and a specificity of 100%.^[8]^ Specimens with an absorbance to Cut-off ratio of ≥ 1.0 was considered as positive. Data collection was done using tablet-based Epi Collect 5 mobile and web-based application was filled for each participant, which included information on age, sex, blood group, symptoms history in past 3 months, complications, contact history, vaccination status and use of mask. Once uploaded, the form was downloaded in Microsoft-Excel data format and merged with registration forms filled at the time of sample collection based on unique identification numbers.

### Data Analysis

The data were extracted in Microsoft excel and analysed in Stata V12. Categorical variables were expressed by proportion whereas the continuous variables were expressed by median, mean and 95% confidence interval. To find the statistical difference, the chi^2^ test was done between categorical variables. The level of significance was taken at 0.05. The corrected estimate was calculated by adjusting the test kit accuracy using the following formula.^[9]^

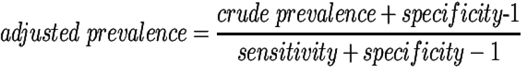

## Result

The data collection period was 15^th^ March to 31^st^ March 2021 (the second wave started April 1^st^, and peak date 20^th^ April) in Delhi urban site, 12^th^ April to 22^nd^ May 2021 (the second wave started April 1^st^, and peak date 5^th^ May) in Delhi rural site (Faridabad, Haryana), 22^nd^ March to 7^th^ May 2021 (the second wave started April 01, and peak date 10 May) in Bhubaneswar site, 22^nd^ April to 10^th^ June 2021 (the second wave started 1^st^ April, and peak date 23^rd^ April) in Gorakhpur site and 26^th^ March to 1^st^ June 2021 (the second wave started April 15^th^, and peak date 23^rd^ May) in Agartala site.

Till 11^th^ June 2021, the number of recruited participants were 1,001 for Delhi urban, 1,059 for Delhi rural, 448 for Gorakhpur rural, 1,000 for Bhubaneswar rural, 1,001 for Agartala rural. The total recruited participants was 4,509. Out of them, the participants of the 2-17 years age group were 92, 189, 165, 146 and 108 for the site of Delhi urban, Delhi rural, Bhubaneswar, Gorakhpur and Agartala respectively aggregating a total of 700. The total number of adults (≥18 years) were 3,809. **(Table No. 1)**. The median age of the analysed participants were 11 years, 12 years, 11 years, 13 years, and 14 years for the site of Delhi urban, Delhi rural, Bhubaneswar, Gorakhpur, and Agartala respectively.

The total number of participants in the 2-17 years age group positive for SARS-CoV-2 antibody was 390/700 (55.7%). The prevalence for the adult participants was 2,421/3,809 (63.5%). The site-wise prevalence in these two age group was almost similar except in Agartala site. **(Table 1)**

**Table 1:** Distribution of participants by age group and COVID-19 sero-positivity at different study sites.

| Variables | Urban Resettlement Colony in South Delhi district<br>n=1001 |  |  | Village at Ballabgarh in district Faridabad, Delhi NCR*<br>n= 1059 |  |  | Bhubaneswar Rural<br>n=1000 |  |  | Agartala Rural<br>n=1001 |  |  | Gorakhpur Rural<br>n=448 |  |  | Total<br>n= 4509 |  |  |  |
| --- | --- | --- | --- | --- | --- | --- | --- | --- | --- | --- | --- | --- | --- | --- | --- | --- | --- | --- | --- |
|  | Partic<br>ipants | Sero-<br>positiv<br>e | Prop<br>sero-<br>positiv<br>e | Parti<br>cipa<br>nts | Sero-<br>positiv<br>e | Prop<br>sero-<br>positiv<br>e | Parti<br>cipa<br>nts | Sero-<br>positiv<br>e | Prop<br>sero-<br>positiv<br>e | Parti<br>cipa<br>nts | Sero-<br>positiv<br>e | Prop<br>sero-<br>positiv<br>e | Parti<br>cipa<br>nts | Sero-<br>positiv<br>e | Prop<br>sero-<br>positiv<br>e | Parti<br>cipa<br>nts | Sero-<br>positiv<br>e | Prop<br>sero-<br>positiv<br>e | Adjust<br>ed<br>preval<br>ence |
|  | n | n | % | n | n | % | n | n | % | n | n | % | n | n | % | n | n | % | % |
| < 18 years | 92 | 68 | 73.9 | 189 | 116 | 61.4 | 165 | 75 | 45.4 | 146 | 45 | 30.8 | 108 | 87 | 80.6 | 700 | 390 | 55.7<br>(52.0-59.4) | 59.0<br>(55.4-62.6) |
| ≥ 18 years | 909 | 680 | 74.8 | 870 | 512 | 58.8 | 835 | 451 | 54.0 | 855 | 475 | 55.6 | 340 | 307 | 90.3 | 3809 | 2421 | 63.5<br>(62.0-65.1) | 67.3<br>(65.8-68.8) |
| **Total** | **1001** | **748** | **74.7** | **1059** | **628** | **59.3** | **1000** | **526** | **52.6** | **1001** | **520** | **51.9** | **448** | **394** | **87.9** | **4509** | **2811** | **62.3**<br>**(60.9-**<br>**63.8)** | **65.9**<br>**(64.6-**<br>**67.4)** |

\*NCR: National capital region

**Table 1A:** Distribution of participants by age group and type of study site.

| Age group | Location |  |  |  |  |  |  |  |
| --- | --- | --- | --- | --- | --- | --- | --- | --- |
|  | Urban |  |  |  | Rural |  |  |  |
|  | Total participants (n) | Total sero-positive (n) | Proportion of sero-positive (%)<br>95% CI | Adjusted prevalence (%)<br>95% CI | Total participants (n) | Total sero-positive (n) | Proportion of sero-positive (%)<br>95% CI | Adjusted prevalence (%)<br>95% CI |
| <18 years | 92 | 68 | 73.9<br>(63.7-82.5) | 78.3 | 608 | 322 | 52.9<br>(49.0-56.9) | 55.9<br>(52.0-59.9) |
| ≥18 years | 909 | 680 | 74.8<br>(71.8-77.6) | 79.2 | 2900 | 1741 | 60.0<br>(58.3-61.8) | 63.5 (61.8-65.3) |
| **Total** | **1001** | **748** | **74.7**<br>**(71.9-77.4)** | **79.1**<br>**(76.5-81.6)** | **3508** | **2063** | **58.8**<br>**(57.2-60.4)** | **62.2**<br>**(60.7-63.9)** |

Irrespective of the age groups, rural sites had lower sero-positivity compared to the urban site (At Delhi). Within the rural sites, children had slightly lower sero-positivity compared to adults. However, this differential prevalence was not observed in the urban site. **(Table 1A)**

The prevalence in children was slightly more among female participants compared to male (58.6% vs 53.0%). However, there was no statistically significant difference (p-value 0.140) in sero-positivity between male and female. Among 700 children aged 2-17 years, 362 (51.7%) were male. The number of participants in the aged 2-4 years were 33 (4.8%), 5-9 years 153 (21.8%), and 10-17 years 512 (73.1%) (**Table 2.)** Children aged 2-4 years and 5-9 year had almost identical sero-positivity rate (42.4% and 43.8%) which was lower than the rate observed for children aged 10-17 years (60.3%). **(Table 2)**

**Table 2:** Distribution of child participants by selected variables and sero-positivity rate.

| Variables | Total (n= 700) |  |  |
| --- | --- | --- | --- |
|  | Participants (n) | Sero-positive (n) | Proportion of sero-positive (%) |
| Male | 362 | 192 | 53.0 |
| Female | 338 | 198 | 58.6 |
| 1-4 years | 33 | 14 | 42.4 |
| 5-9 years | 153 | 67 | 43.8 |
| 10-17 years | 512 | 309 | 60.3 |
| Symptomatic | 131 | 90 | 68.7 |
| Asymptomatic | 569 | 300 | 52.7 |
| **Total** | **700** | **390** | **55.7** |

**Table 3:**
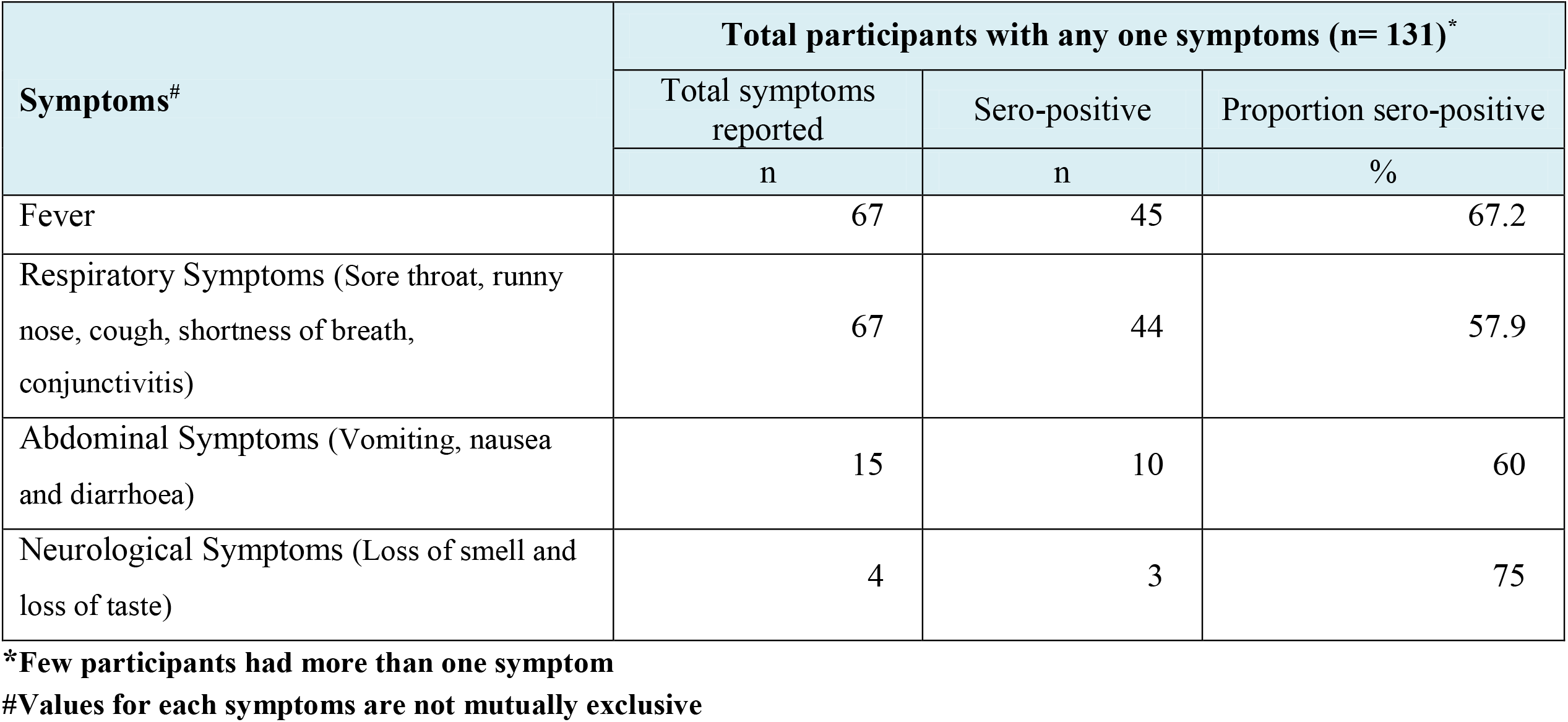
Distribution of participants <18 years by symptoms and sero-positivity rate.

## Discussion

There was a slightly higher sero-positivity rate observed among female children. This finding was in contrast to the meta-analysis where it was shown that the prevalence is higher in men.^[10]^ This may be a chance finding due to small number of data available at the time of midterm analysis. The higher seropositivity rate in children aged 10-17 years may be reflective of their higher mobility and independence compared to the younger children. As reported in the literature, a large proportion of children (50.9%) had asymptomatic COVID-19 infection.^[11]^

In India, sero-prevalence among children and younger age group were estimated as a part of a larger nationwide survey on adult age group. The second nationwide sero-prevalence study done in August-September 2020 had reported 9.0% seropositive among 3,021 children aged 10-17 years.^[12]^ while in our study it is 60.3%. One hospital-based study in Chennai had reported 19.6% prevalence in the age group of 1 month to 17 years.^[13]^

### Delhi Urban

During the first wave of the pandemic in India, the worst affected areas were the large urban areas, including Delhi. We collected the data during the second fortnight of March 2021. This was the time when the first wave was subsiding and the second wave had not yet started. Results show that a large majority of the population had already been infected by the time we conducted the study at Delhi urban site which belongs to lower and middle socioeconomic strata population and very congested neighbourhood. The obliteration of any difference in sero-positivity rate between children and adult suggests that as the disease become generalized, it affects all age groups equally.

We found that sero-positivity rate in our study was higher (74.7%) than the fifth sero survey (conducted in January 2021) which reported an overall 56.1% for Delhi and 62.8% for South Delhi district.^[14,15]^ However, information on prevalence in the child and young age group was lacking.

**Rur**al -We had included four rural sites. Two of the sites (Bhubaneswar and Agartala) were state capital, one site (Faridabad under the rural site of AIIMS, Delhi) was in National Capital Region and one site (Gorakhpur) was a major transit point for surface transportation to another country (Nepal). Thus these sites were more vulnerable to a pandemic. The data were collected during the second wave. Gorakhpur site was the worst affected (sero-positivity rate of 87.9%) while Faridabad was the least affected (sero-positivity rate 58.8%). The data collection of all the site was done just before the second wave hit the country except the site of Gorakhpur. This may be the reason for the highest prevalence at the Gorakhpur site among all the sites. Overall, more than half (62.3%) of those surveyed showed evidence of past infection. Agartala site included some tribal population as well. In general tribal population had lower mobility resulting in lower vulnerability to COVID19 infection. This might explain the comparatively low prevalence in children at this site.

In a rapidly evolving pandemic, individuals who have been recently infected (< 14 days) may not have developed antibodies. They would have been reported negative in sero-survey. Hence, our findings are likely to be an underestimate.

We observe that children had a slightly lower sero-positivity rate compared to adults (55.7% vs 63.5%). These findings are similar to the previously reported evidence which found that children are less affected than the adult age group.^[16,17]^ During the pandemic schools were closed and children were more likely to have remained indoors compared to adults. For children, the source of infections is likely to be the household adults who brought the infection from outside during livelihood activities. Hence, we can expect some lag in sero-positivity among children. We are not sure if children produce the same level of antibodies as adults when infected. If children produce a lower level of antibodies that might not be detectable by the existing laboratory tests, then the observed sero-positivity rate would be a reflection of the laboratory tests proficiency rather than any true difference between children and adult infection rates. Overall, the results suggest that children and adults are equally susceptible to COVID-19 infection.

### Strength

This study included participants from four different states representing different geographical locations of India. We, for the first time, provide sero-prevalence estimate for children aged 2-17 years. Data from both urban slum area, rural area and some tribal population at one site, further increases the generalizability. By including a large number of clusters (25) within each of the study site makes our findings more representative.

### Limitation

The sampling method was cluster sampling. In Delhi urban, we had purposively selected an urban resettlement colony. This area had high population density inhabited mostly by lower socio-economic population. Therefore, it is not representative of Delhi urban population as a whole. At all other sites, clusters were selected randomly. Participation was voluntary in our study. Secondly, we did not perform neutralizing antibody assay for all these samples that would have helped to interpret the significance of sero-positivity.

### Conclusion

SARS-CoV-2 sero-positivity rate among children was high and comparable to the adult population. Hence, it is unlikely that any future third wave by prevailing COVID-19 variant would disproportionately affect children two years or older.

## Data Availability

yes available with us and WHO

## Declarations

### Ethical Approval and Consent to participate

The study received ethical clearance from all five participating institutes (Letter No. For AIIMS, New Delhi: **IEC-959/04.09.2020**, AIIMS Bhubaneswar: **T/EMF/CM&FM/20/44**, JIPMER Puducherry: **JIP/IEC/2020/248**, AIIMS Gorakhpur: **IHEC/AIIMS-GKP/BMR/01/22**, Agartala: **F.4(5-234)/AGMC/ACADEMIC/IEC MEETING**). Written informed consent, assent and parental consent for participants under the legal age of giving consent was taken from all the participants as per ICMR guidelines.

### Funding

This work was supported by a research grant (Ref No: **2020/1085497**, Purchase Order: **202630166**) from the WHO Country Office, New Delhi 110016, India.

### Conflicts of interest

The authors declare no conflict of interests regarding the publication of this paper.

## Acknowledgement

We thank the WHO Country Office, India team particularly Dr Mohammad Ahmad (National Professional Officer, WHO) and Dr Anisur Rahman (Health Emergencies and Research Officer, WHO) for continuous support. Special thanks to the participating subjects who allowed us to investigate the extent of infection, as determined by seropositivity in the general population, in which COVID-19 virus infection has been reported.

---

**Figure 1:**
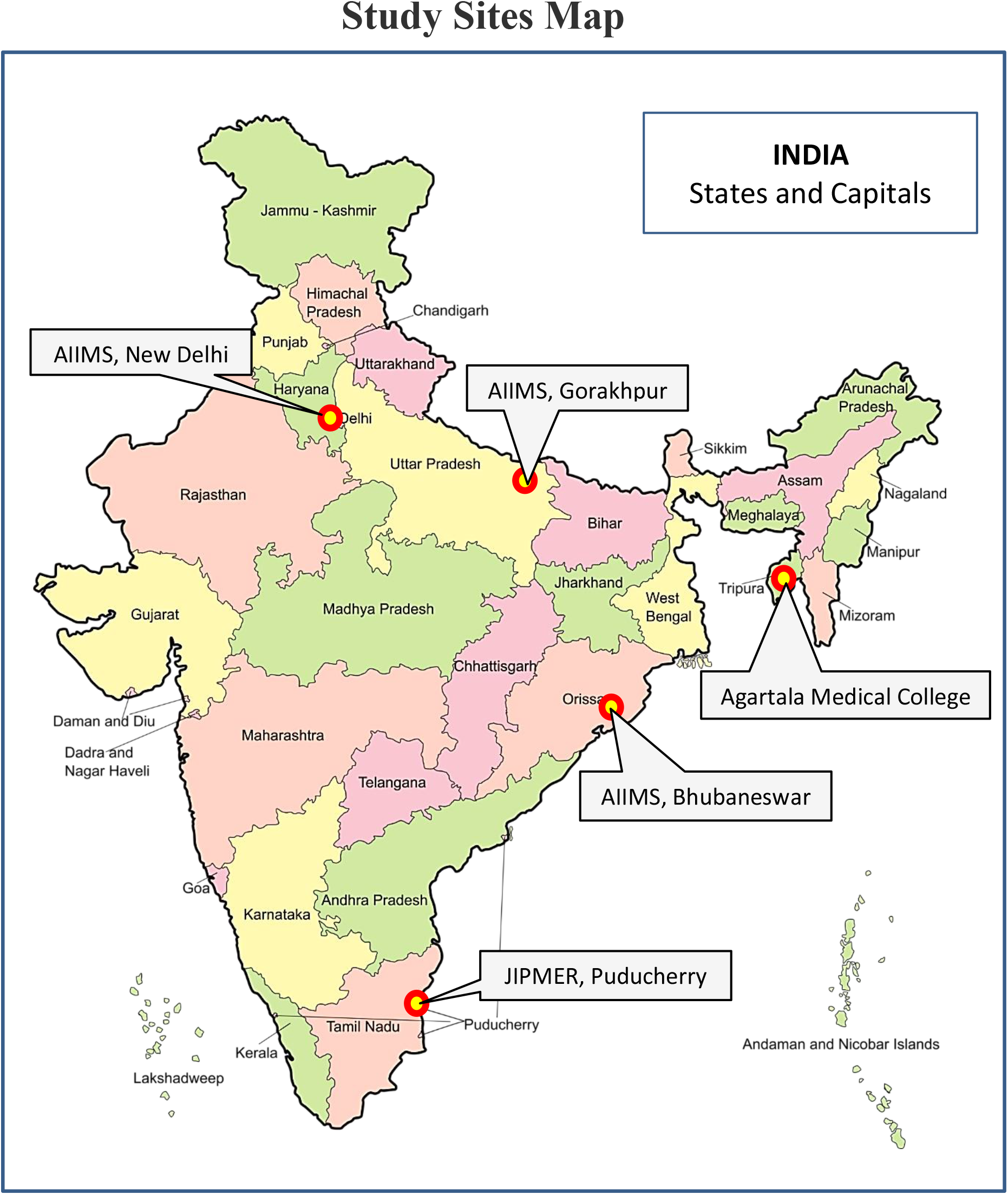
The geographical location of the study sites in India Partnering Institutes 1. All India Institute of Medical Sciences, New Delhi 110029, India 2. All India Institute of Medical Sciences, Bhubaneshwar, Odisha- 751019, India 3. All India Institute of Medical Sciences, Gorakhpur- 273008, India 4. Jawaharlal Institute of Postgraduate Medical Education and Research, Puducherry- 605006, India 5. Agartala Government Medical College, Agartala- 799006, India 6. Translational Health Science and Technology Institute, Faridabad-121001, India

## Notes

### Competing Interest Statement

The authors have declared no competing interest.

### Author Declarations

The study received ethical clearance from all five participating institutes (Letter No. For AIIMS, New Delhi: IEC-959/04.09.2020, AIIMS Bhubaneswar: T/EMF/CM&FM/20/44, JIPMER Puducherry: JIP/IEC/2020/248, AIIMS Gorakhpur: IHEC/AIIMS-GKP/BMR/01/22, Agartala: F.4(5-234)/AGMC/ACADEMIC/IEC MEETING). Written informed consent, assent and parental consent for participants under the legal age of giving consent was taken from all the participants as per ICMR guidelines.

